## Supplementary Dataset 1 for "Plasma p-tau212: antemortem diagnostic performance and prediction of autopsy verification of Alzheimer’s disease neuropathology"

**Supplementary Data Table 1. Peptides detected in the antibody specificity experiment by IP-MS. Only peptides with p-tau212 present were detected.**

| **Extended-peptide standard^a^** | **Extended peptide sequence^b^** | ***m/z*** | **Charge** | **Detected peptide** |
| --- | --- | --- | --- | --- |
| 170-195-pT181 | RIPA(K)TPPAP(K)[T]PPSSGEPP(K)SGDRS | 561.949 | 3+ |  |
| 190-214-pS202 | KSGDRSGYSSPG[S]PGTPGS(R)SRTPS | 742.345 | 2+ |  |
| 190-214-pT205 | KSGDRSGYSSPGSPG[T]PGS(R)SRTPS | 742.345 | 2+ |  |
| 190-214-pS202+pT205 | KSGDRSGYSSPG[S]PG[T]PGS(R)SRTPS | 782.291 | 2+ |  |
| 205-229-pT212 | TPGSRSR[T]PSLPTPPT(R)EP(K)KVAVV | 587.972 | 3+ | X |
| 205-229-pS214 | TPGSRSRTP[S]LPTPPT(R)EP(K)KVAVV | 587.972 | 3+ |  |
| 205-229-pT217 | TPGSRSRTPSLP[T]PPT(R)EP(K)KVAVV | 587.972 | 3+ |  |
| 205-229-pT212+pT217 | TPGSRSR[T]PSLP[T]PPT(R)EP(K)KVAVV | 614.627 | 3+ | X |
| 220-245-pT231 | TREPKKVAVV(R)[T]PP(K)SPSSA(K)SRLQT | 586.668 | 3+ |  |

^a^ Numbering according to tau 2N4R

^b^ [X] amino acid with phosphate group, (X) isotope labelled amino acid; [^13^C_6_,^15^N_2_-Lys] or [^13^C_6_,^15^N_4_-Arg]; black letters are detected peptide after trypsinization, and red letters are extended sequence parts

**Supplementary Data Table 2. Neurofibrillary tangles and neuropil threads count in slices of AD brains.**

| **Tangles Count** | **p212** | **AT8** |  | **Tangles**  **Count** | **p212** | **p217** |
| --- | --- | --- | --- | --- | --- | --- |
| **AD1** | 17 | 17 |  | **AD1** | 6 | 6 |
| **AD2** | 10 | 10 |  | **AD2** | 7 | 7 |
| **AD3** | 39 | 36 |  | **AD3** | 18 | 18 |
| **Threads Count** | **p212** | **AT8** |  | **Threads Count** | **p212** | **p217** |
| **AD1** | 385 | 415 |  | **AD1** | 355 | 331 |
| **AD2** | 154 | 155 |  | **AD2** | 336 | 326 |
| **AD3** | 301 | 243 |  | **AD3** | 363 | 359 |

**Supplementary Data Table 3. The %parallelism values for each subsequent dilution factor for Extended Data Figure 1.**

| **Plasma parallelism** | | | |
| --- | --- | --- | --- |
| **Dilution/Sample** | **Sample 1** | **Sample 2** | **Sample 3** |
| 2-fold | 92.3 | 106.0 | 121.7 |
| 4-fold | 78.6 | 84.0 | 118.7 |
| **CSF parallelism** | | | |
| **Sample/%Parallelism** | **Sample 1** | **Sample 2** | **Sample 3** |
| 32-fold | 100.7 | 87.2 | 81.7 |
| 64-fold | 97.4 | 87.5 | 91.0 |

**Supplementary Data Table 4. The coefficients of variation (%CV) Internal Quality Control samples**.

| **Cohort** | **Internal Quality control (iQC) sample** | **% CV** | | | |
| --- | --- | --- | --- | --- | --- |
|  |  | **P-tau181** | **P-tau212** | **P-tau217** | **P-tau231** |
| Gothenburg | iQC 1 | 0.2% | 3.2% | - | 0.6% |
|  | iQC 2 | 5.7% | 3.6% | - | 12.0% |
| Polish - CSF | iQC 1 | - | 3.3% within run and 7.1% between-run | - | - |
|  | iQC 2 | - | 9.6% within run and 7.1% between-run | - | - |
| Polish - plasma | iQC 1 | - | 8.3% within run and 19.0% between-run | - | - |
|  | iQC 2 | - | 19.0% within run and 19.9% between-run | - | - |
| BLSA-Neuropathology | iQC 1 | 8.1% | 1.0% | - | 1.3% |
|  | iQC 2 | 4.6% | 5.2% |  | 3.8% |
| UCSD-Neuropathology | iQC 1 | 2.8% | 5.7% | 1.6% | 1.1% |
|  | iQC 2 | 2.7% | 4.1% | 14.7% | 2.5% |
| Slovenia | iQC 1 | - | 13.7% within run and 13.7% between-run | 4.5% within run and 17.8% between-run | - |
|  | iQC 2 | - | 14.7% within run and 14.7% between-run | 4.8% within run and 20.5% between-run | - |

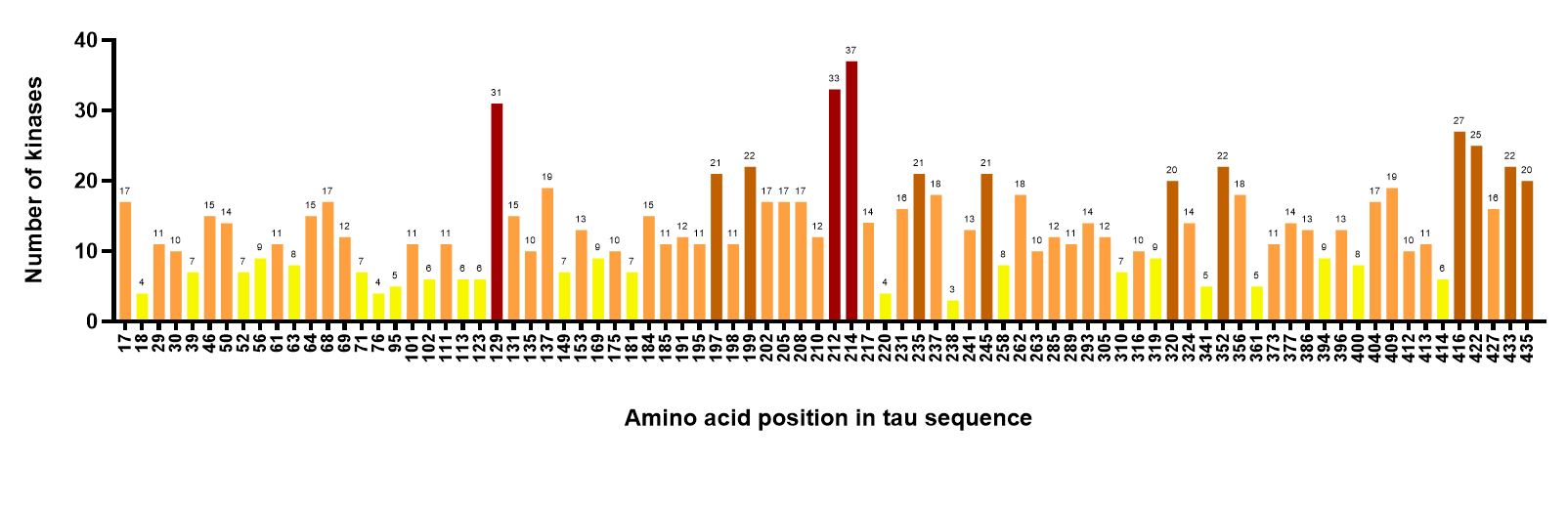
**Supplementary Data Figure 1. Computational prediction of epitopes in full-length tau-441 protein phosphorylatable by unique kinases.**

The figure shows the number of different kinases that are predicted to phosphorylate single epitopes in human tau protein 2N4R (Uniprot ID: P10636-8) created by the computational tool GPS 5.0. The epitopes are arranged in the order of amino acid positions in tau protein. The colour coding corresponds to the number of predicted kinases. The numbers at the top of each bar indicates the total number of predicted kinases.

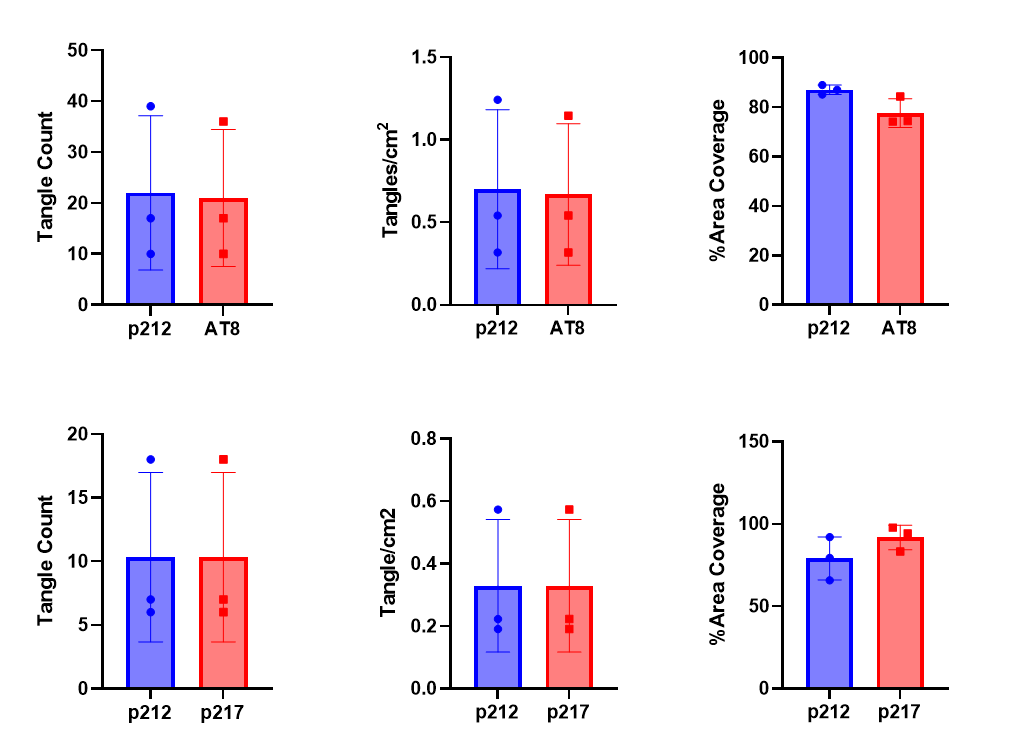

**Supplementary Data Figure 2. Bar plot representation of tangle count and %area of tangles covered by each antibody in the immunofluorescent staining experiments.**

Three independent AD brains were evaluated. In each plot, a single dot refers to data from one of these individuals. The data are presented as mean values and the vertical bars show standard deviation.

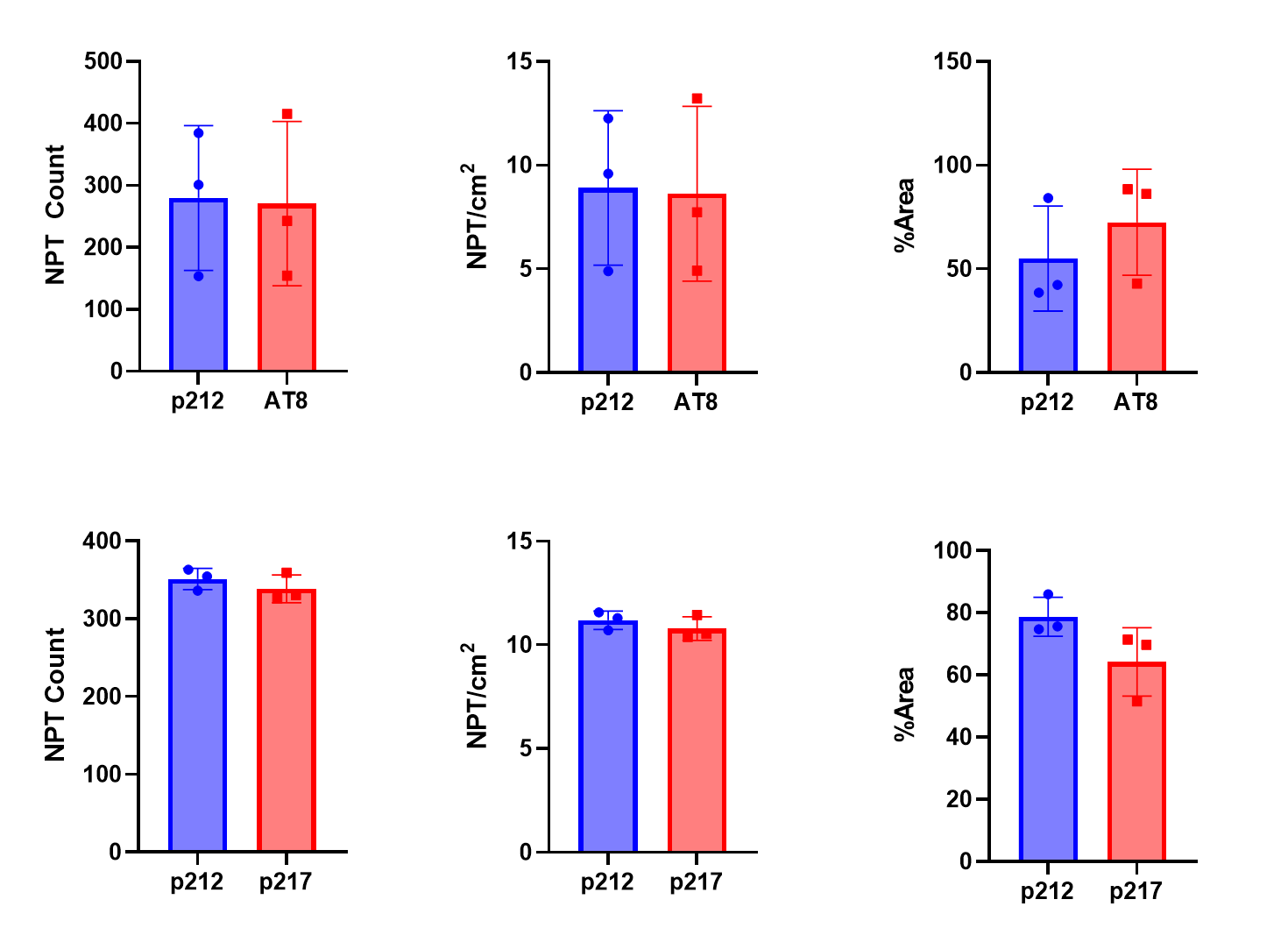

**Supplementary Data Figure 3. Bar plot representation of neuropil threads count and %area of neuropil threads covered by each antibody in the immunofluorescent staining experiments.**

Three independent AD brains were evaluated. In each plot, a single dot refers to data from one of these individuals. The data are presented as mean values and the vertical bars show standard deviation.

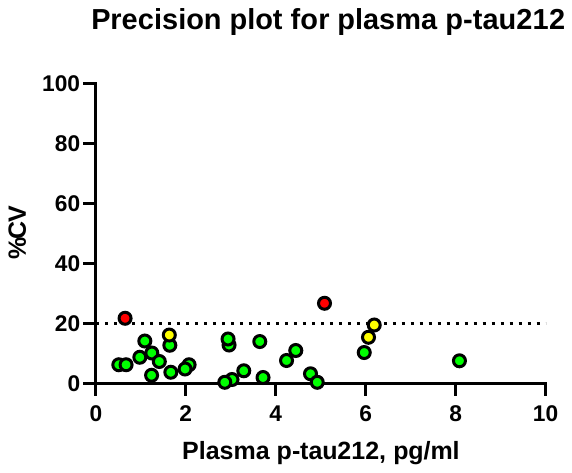

**Supplementary Data Figure 4. Coefficients of variation for p-tau212 in de-identified plasma samples.**

The figure shows %CV values for n=30 de-identified plasma samples ran in duplicates. %CV’s below 15% were marked as green dots; %CV’s above 15% and below 20% were marked as yellow dots and for duplicates that had %CV over 20% dots were marked in red colour.

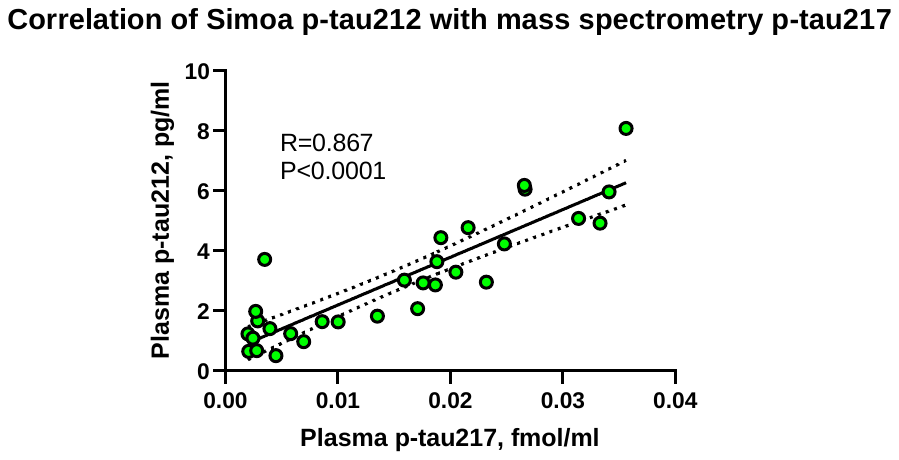

**Supplementary Data Figure 5. Spearman Correlation for IP-MS plasma p-tau217 and Simoa plasma p-tau212**

De-identified plasma samples were run both for the p-tau212 Simoa immunoassay and the IP-MS p-tau217 method. Spearman correlation of both measurements is presented on the graph.

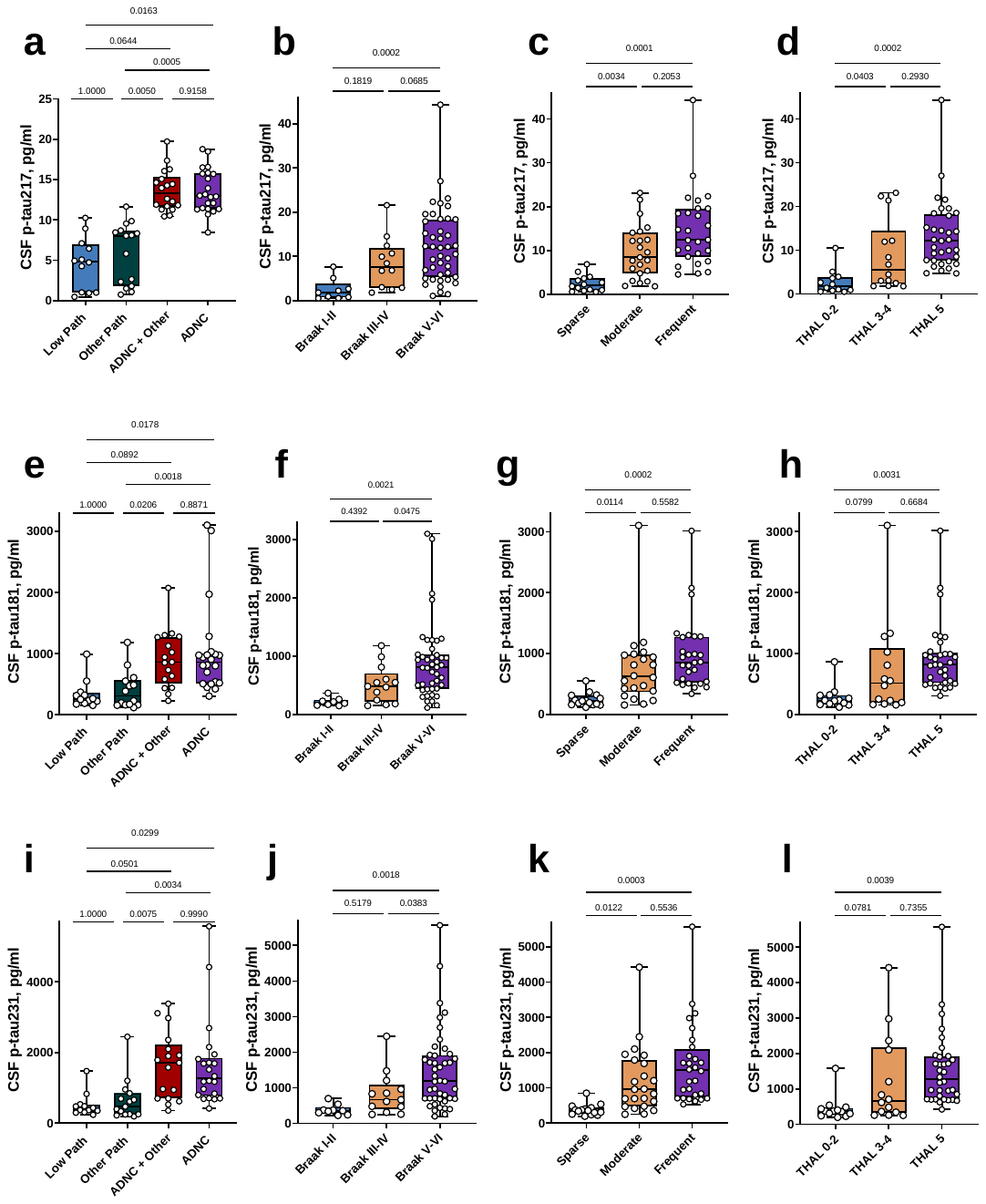

**Supplementary Data Figure 6. Clinical performance of CSF p-tau217, p-tau181 and p-tau231 in the UCSD-Neuropathology cohort.**

The figure exhibits boxplots of the measured levels of CSF p-tau181, p-tau217, p-tau231 in UCSD-Neuropathology cohort.
